# A COVID-19 Reopening Readiness Index: The Key to Opening up the Economy

**DOI:** 10.1101/2020.05.22.20110577

**Authors:** Eunju Suh, Mahdi Alhaery

## Abstract

With respect to reopening the economy as a result of the COVID-19 restrictions, governmental response and messaging have been inconsistent, and policies have varied by state as this is a uniquely polarizing topic. Considering the urgent need to return to normalcy, a method was devised to determine the degree of progress any state has made in containing the spread of COVID-19. Using various measures for each state including mortality, hospitalizations, testing capacity, number of infections and infection rate has allowed for the creation of a composite COVID -19 Reopening Readiness Index. This index can serve as a comprehensive reliable and simple-to-use metric to assess the level of containment in any state and to determine the level of risk in further opening. As states struggle to contain the outbreak and at the same time face great pressure in resuming economic activity, an index that provides a data-driven and objective insight is urgently needed.

**Background:** We are in the midst of a once-in-a-lifetime pandemic. All levels of society and governments are working together to “flatten the curve” of the infection and slow the spread of COVID-19. The universal goal is to mitigate its adverse effects on everyday life across the globe and to reduce the number of fatalities. While a vaccine is being developed, the aim is to limit the number of hospitalizations as not to overwhelm healthcare systems in any given city or country. It is well documented that certain regions and localities are more affected than others. It is imperative that containment efforts utilize state and local data at their disposal to understand the readiness of any given area prior to opening its economy, and the level of restrictions that are needed.

## Introduction

### Reopening Guidelines

There is universal agreement on the need to resume normal economic activity at some point in the near future. The effect of the pandemic on world economies has been devastating. Additionally, there is a need to have plans in places to closely monitor the effect of easing the imposed restrictions. In the United States, the federal administration under President Trump has recently revealed a three-phase plan with guidelines to reopen the American economy. The guidelines were designed to help state and local officials reopen their economies while slowing the spread of COVID-19 and saving lives. These recommendations are as follows:

- SYMPTOMS

- Downward trajectory of influenza-like illnesses (ILI) reported within a 14-day period AND
- Downward trajectory of COVID-19-like syndromic cases reported within a 14-day period
- CASES

- Downward trajectory of documented cases within a 14-day period OR
- Downward trajectory of positive tests as a percent of total tests within a 14-day period (flat or increasing volume of tests)
- HOSPITALS

- Treat all patients without crisis care AND
- Robust testing program in place for at-risk healthcare workers, including emerging antibody testing

(Source: https://www.whitehouse.gov/openingamerica/)

### Potential Issues with the Reopening Metrics

While the federal guidelines note the data-driven strategy to move forward to the next phase of reopening, it leaves reopening decisions to state and local officials by giving flexibility to tailor the application of the aforementioned criteria to local circumstances and needs. Most states appear ready to proceed with the phased plan, and some formed their own regional pacts to plan the region’s reopening (Fink, 2020). When it comes to the criteria for determining when to ease social distancing and other restrictions, to the best of the authors’ knowledge, states have adopted their own internal metrics to monitor progress towards satisfying these criteria.

States typically report key COVID-19 metrics such as the number of deaths, infections, hospitalized cases and tests given. Along with these metrics, some states adopted additional criteria. For example, the state of Colorado monitors the two-week cumulative incidence rate which indicates new cases reported in the past two weeks per 100,000 people. The incident rate can be categorized into one of four groups depending on whether it is above or below the cut off value: low if the number of new cases are 10 or fewer per 100,000 people in the past two weeks, moderate if the number of new cases are between 10 and 50 for the same timeframe, moderately high for 50 - 100 new cases, and high for 100 plus cases. The state also reports the trajectory of the three-day moving average daily incidence per 100,000 to gauge whether the incident growth is low, elevated, reached plateau or declining (CDPHE, 2020). Some other states report additional metrics which are not necessarily available in other states. For example, the state of North Carolina monitors the number of contact tracers and personal protective equipment (PPE) for critical healthcare and frontline workers as well as the number of COVID-like illnesses, confirmed COVID-19 cases and positive tests as a percentage of total tests (NCDHHS, 2020). Despite various methods and metrics, there appears to be no universal criteria among the states with respect to tracking the success of containment.

In addition to the lack of consistency in the methods and metrics tracking COVID-19s impact, the political divide in reopening a state is notable. Differences in reopening approaches and timelines are easily observed between Republican governors and Democrat governors (Blackmon, 2020). Of the 24 states partially reopened as of May 5th, 2020, 21 voted for President Trump in 2016 (Brewster, 2020). With the mounting unemployment rate observed across the U.S., and the nationwide protests demanding governors to ease the restrictions, the process of opening the economy can be easily influenced by political and economic pressure rather than objective criteria. Medical experts and pundits caution that opening up the economy and easing restrictions prematurely can cause a rebound of COVID-19 cases (Fink, 2020).

## The Need for an Index

Many governments have proposed a phased approach to easing the restrictions. These phases will depend upon several factors. The chosen factors vary by state and are somewhat difficult to quantify. A lack of objective criteria and monitoring towards satisfying the criteria can lead to another COVID-19 outbreak and business closures. In an effort to strengthen the readiness of states to reopen their economies and move to the next phase of reopening, this study proposes a COVID-19 Reopening Readiness Index by which all states would be treated equally as to the suitability of opening the economy. The index is based on five broad categories: COVID-19 cases, infection rate, hospitalization, testing and mortality. These criteria are indicative of the spread of an infectious disease and success in combating it. The developed metric distills all of the information into one easy-to-use and easy-to-understand universal index to determine the level of restrictions needed.

The index tracks each state’s readiness to move to the next phase by addressing the guidelines provided by White House. This index provides a universal and unbiased way of determining whether an economy should open and the needed level of restriction. Furthermore, it indicates the degree of success any given government has had and will signal if any given state can further ease the restrictions or if there is a need to revert to a more restrictive plan.

## Methodology

### Indicators of Reopening Readiness

Based on the criteria suggested by the federal government and a few other state governments, we have identified available tracking data to create an index that will determine the degree of progress any state has made in containing the outbreak of the virus. First, a sub-index was created for each of the identified indicators related to COVID-19: the number of new daily cases, the number of hospitalized patients, death counts, tests given, and positive tests as a percent of total tests at a state level. These indexes are calculated based on the three-day rolling averages of the key metrics to minimize irregular fluctuation in the daily data and still provide easily identifiable trends over time. Next, all of the sub-indexes were combined to produce a composite index. A composite index provides a comprehensive picture of the effectiveness of the government’s restriction in limiting the spread of the COVID-19 and success in combatting it. The following table presents the key indicators associated with the sub-indexes used to create a composite COVID-19 Reopening Readiness Index.

**Table 1.** Key Indicators and Indexes of COVID-19 Reopening Readiness

| Sub-Index | Key Indicator | Sign of Progress |
| --- | --- | --- |
| Cases | Rolling three-day averages of the number of daily positive cases | A consistent decrease in the Cases Index number |
| Infection Rate | Rolling three-day averages of the number of daily positive tests as a percent of total tests | A consistent decrease in the Infection Rate Index number |
| Mortality | Rolling three-day averages of the number of daily death counts | A consistent decrease in the Mortality Index number |
| Hospitalization | Rolling three-day averages of the number of daily hospitalized cases | A consistent decrease in the Hospitalization Index number |
| Testing Capacity | Rolling three-day averages of the number of daily tests | A constant or an increase in the Testing Capacity Index number showing an increase in tests administered for population |

Three-day rolling averages were used to produce a smoother curve and accommodate small or missing counts and fluctuations found in the daily values because a small change in the numerator can result in a large change in the index value. The following section presents the steps in calculating the sub-indexes and the composite COVID-19 Reopening Readiness Index.

### Steps for Calculating Indexes

Data from *The COVID Tracking Project* (COVID Tracking, 2020) was used to create the indicators and the associated sub-indexes.

Step 1. Generate the three-day rolling average of daily counts

Step 2. Calculate the ratio of the current day’s count (the average of counts for the current and past two days) divided by the three-day average prior to the current day

Step 3. Repeats Step 1 and Step 2 for each day and indicator

Step 4. Create sub-indexes by normalizing the ratios using the following formula:

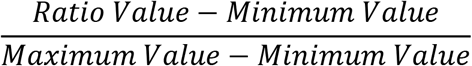

Normalization enables comparisons of data across indicators by removing the units of measurement for data. Abnormal and extreme values found in the data set, which appear to be due to irregular updates, were either removed or replaced with the average plus two standard deviations of a sample during the evaluation period. As outliers can skew the data and result in misleading interpretation, they are replaced with the above-mentioned values. Approximately 95% of the data falls within two standard deviations of the mean. Minimum and maximum values were identified in the data set for each indicator. Finally, aggregating each of the sub-indexes together allows for the creation of the COVID-19 Reopening Readiness Index which is a composite index useful for measuring progress in containing the outbreak of the virus.

Step 5. Calculate the arithmetic mean of the sub-indicates to generate a composite index

The COVID-19 Reopening Readiness Index is equal-weighed not to overemphasize one index over the others. For example, a state may experience a decline in death counts but an increase in hospitalization cases. Another scenario could be an increase in the number of infection cases but a decrease in the infection rate possibly because of the increase in the testing capacity. Giving equal weight to each of the indexes prevents using the criterion that only meet expectations to justify reopening.

With respect to the specific sub-index representing testing capacity, the value 1minus sub-index was used in the calculation of a composite index such that the index value can be interpreted in the same way as other sub-indexes. As data availability varies by state, a composite index was calculated based on the sub-indexes derived from the available data. Where data was not found in the data set, the composite index would be the combination of only the available sub-indexes (See Appendix for more details).

## Results

### COVID-19 Reopening Readiness Index

The composite index allows users to compare a given state’s performance over time. Its value ranges from 0 to 1 where the higher values correspond to a higher level of risk in reopening the state. Table 2 shows the COVID-19 Reopening Readiness Index values derived from the multiple COVID-19 sub-indexes by state.

**Table 2.**
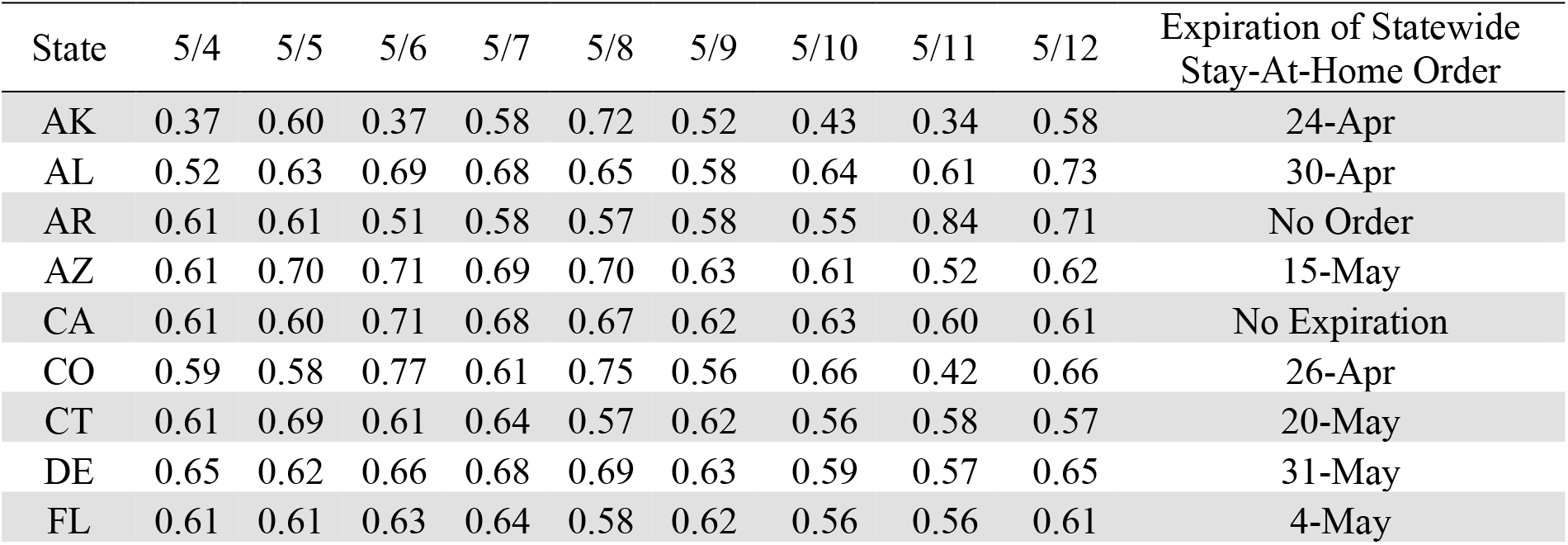
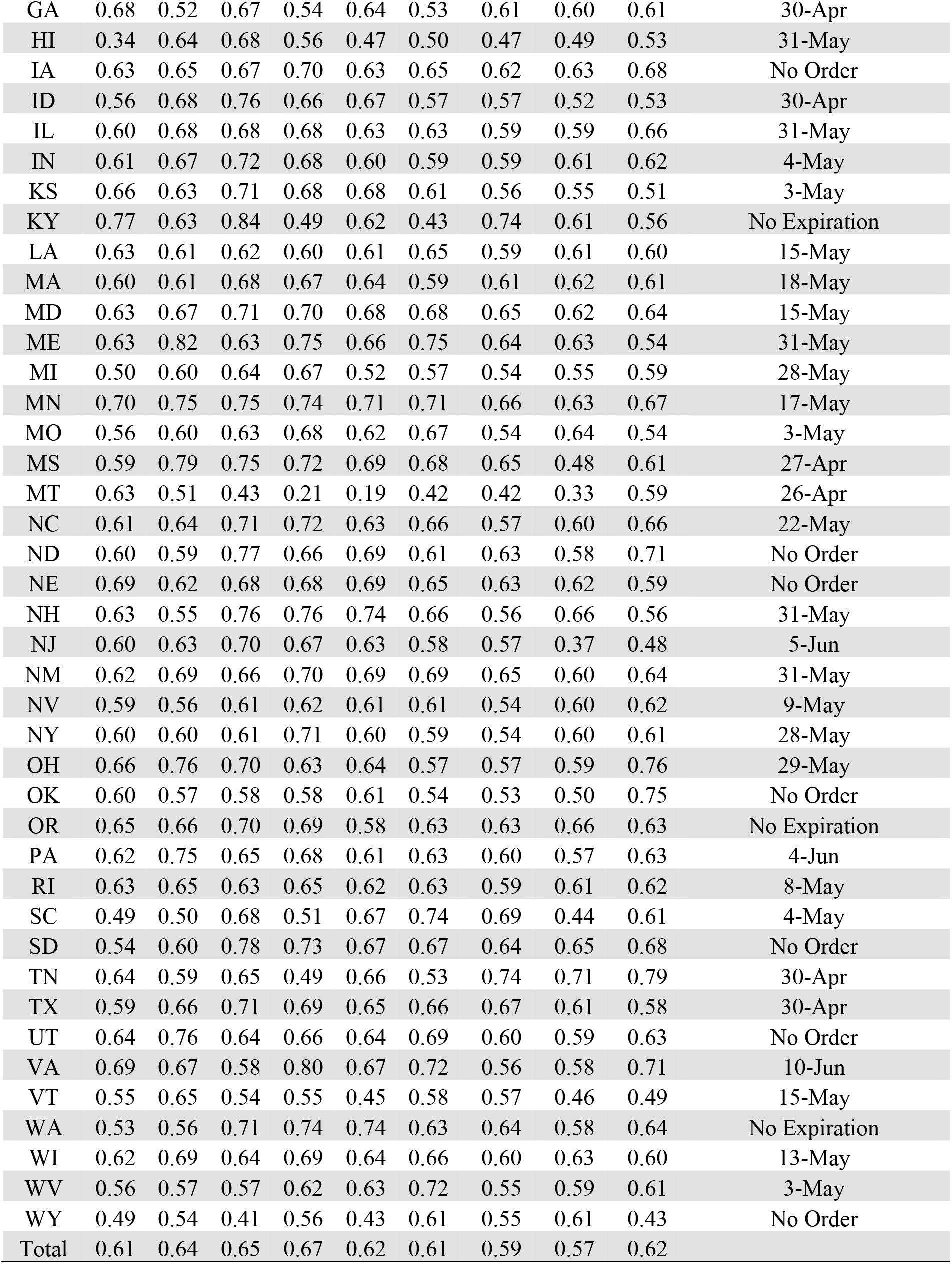
Daily Index of COVID-19 Reopening Readiness

| State | 5/4 | 5/5 | 5/6 | 5/7 | 5/8 | 5/9 | 5/10 | 5/11 | 5/12 | Expiration of Statewide Stay-At-Home Order |
| --- | --- | --- | --- | --- | --- | --- | --- | --- | --- | --- |
| AK | 0.37 | 0.60 | 0.37 | 0.58 | 0.72 | 0.52 | 0.43 | 0.34 | 0.58 | 24-Apr |
| AL | 0.52 | 0.63 | 0.69 | 0.68 | 0.65 | 0.58 | 0.64 | 0.61 | 0.73 | 30-Apr |
| AR | 0.61 | 0.61 | 0.51 | 0.58 | 0.57 | 0.58 | 0.55 | 0.84 | 0.71 | No Order |
| AZ | 0.61 | 0.70 | 0.71 | 0.69 | 0.70 | 0.63 | 0.61 | 0.52 | 0.62 | 15-May |
| CA | 0.61 | 0.60 | 0.71 | 0.68 | 0.67 | 0.62 | 0.63 | 0.60 | 0.61 | No Expiration |
| CO | 0.59 | 0.58 | 0.77 | 0.61 | 0.75 | 0.56 | 0.66 | 0.42 | 0.66 | 26-Apr |
| CT | 0.61 | 0.69 | 0.61 | 0.64 | 0.57 | 0.62 | 0.56 | 0.58 | 0.57 | 20-May |
| DE | 0.65 | 0.62 | 0.66 | 0.68 | 0.69 | 0.63 | 0.59 | 0.57 | 0.65 | 31-May |
| FL | 0.61 | 0.61 | 0.63 | 0.64 | 0.58 | 0.62 | 0.56 | 0.56 | 0.61 | 4-May |
| GA | 0.68 | 0.52 | 0.67 | 0.54 | 0.64 | 0.53 | 0.61 | 0.60 | 0.61 | 30-Apr |
| HI | 0.34 | 0.64 | 0.68 | 0.56 | 0.47 | 0.50 | 0.47 | 0.49 | 0.53 | 31-May |
| IA | 0.63 | 0.65 | 0.67 | 0.70 | 0.63 | 0.65 | 0.62 | 0.63 | 0.68 | No Order |
| ID | 0.56 | 0.68 | 0.76 | 0.66 | 0.67 | 0.57 | 0.57 | 0.52 | 0.53 | 30-Apr |
| IL | 0.60 | 0.68 | 0.68 | 0.68 | 0.63 | 0.63 | 0.59 | 0.59 | 0.66 | 31-May |
| IN | 0.61 | 0.67 | 0.72 | 0.68 | 0.60 | 0.59 | 0.59 | 0.61 | 0.62 | 4-May |
| KS | 0.66 | 0.63 | 0.71 | 0.68 | 0.68 | 0.61 | 0.56 | 0.55 | 0.51 | 3-May |
| KY | 0.77 | 0.63 | 0.84 | 0.49 | 0.62 | 0.43 | 0.74 | 0.61 | 0.56 | No Expiration |
| LA | 0.63 | 0.61 | 0.62 | 0.60 | 0.61 | 0.65 | 0.59 | 0.61 | 0.60 | 15-May |
| MA | 0.60 | 0.61 | 0.68 | 0.67 | 0.64 | 0.59 | 0.61 | 0.62 | 0.61 | 18-May |
| MD | 0.63 | 0.67 | 0.71 | 0.70 | 0.68 | 0.68 | 0.65 | 0.62 | 0.64 | 15-May |
| ME | 0.63 | 0.82 | 0.63 | 0.75 | 0.66 | 0.75 | 0.64 | 0.63 | 0.54 | 31-May |
| MI | 0.50 | 0.60 | 0.64 | 0.67 | 0.52 | 0.57 | 0.54 | 0.55 | 0.59 | 28-May |
| MN | 0.70 | 0.75 | 0.75 | 0.74 | 0.71 | 0.71 | 0.66 | 0.63 | 0.67 | 17-May |
| MO | 0.56 | 0.60 | 0.63 | 0.68 | 0.62 | 0.67 | 0.54 | 0.64 | 0.54 | 3-May |
| MS | 0.59 | 0.79 | 0.75 | 0.72 | 0.69 | 0.68 | 0.65 | 0.48 | 0.61 | 27-Apr |
| MT | 0.63 | 0.51 | 0.43 | 0.21 | 0.19 | 0.42 | 0.42 | 0.33 | 0.59 | 26-Apr |
| NC | 0.61 | 0.64 | 0.71 | 0.72 | 0.63 | 0.66 | 0.57 | 0.60 | 0.66 | 22-May |
| ND | 0.60 | 0.59 | 0.77 | 0.66 | 0.69 | 0.61 | 0.63 | 0.58 | 0.71 | No Order |
| NE | 0.69 | 0.62 | 0.68 | 0.68 | 0.69 | 0.65 | 0.63 | 0.62 | 0.59 | No Order |
| NH | 0.63 | 0.55 | 0.76 | 0.76 | 0.74 | 0.66 | 0.56 | 0.66 | 0.56 | 31-May |
| NJ | 0.60 | 0.63 | 0.70 | 0.67 | 0.63 | 0.58 | 0.57 | 0.37 | 0.48 | 5-Jun |
| NM | 0.62 | 0.69 | 0.66 | 0.70 | 0.69 | 0.69 | 0.65 | 0.60 | 0.64 | 31-May |
| NV | 0.59 | 0.56 | 0.61 | 0.62 | 0.61 | 0.61 | 0.54 | 0.60 | 0.62 | 9-May |
| NY | 0.60 | 0.60 | 0.61 | 0.71 | 0.60 | 0.59 | 0.54 | 0.60 | 0.61 | 28-May |
| OH | 0.66 | 0.76 | 0.70 | 0.63 | 0.64 | 0.57 | 0.57 | 0.59 | 0.76 | 29-May |
| OK | 0.60 | 0.57 | 0.58 | 0.58 | 0.61 | 0.54 | 0.53 | 0.50 | 0.75 | No Order |
| OR | 0.65 | 0.66 | 0.70 | 0.69 | 0.58 | 0.63 | 0.63 | 0.66 | 0.63 | No Expiration |
| PA | 0.62 | 0.75 | 0.65 | 0.68 | 0.61 | 0.63 | 0.60 | 0.57 | 0.63 | 4-Jun |
| RI | 0.63 | 0.65 | 0.63 | 0.65 | 0.62 | 0.63 | 0.59 | 0.61 | 0.62 | 8-May |
| SC | 0.49 | 0.50 | 0.68 | 0.51 | 0.67 | 0.74 | 0.69 | 0.44 | 0.61 | 4-May |
| SD | 0.54 | 0.60 | 0.78 | 0.73 | 0.67 | 0.67 | 0.64 | 0.65 | 0.68 | No Order |
| TN | 0.64 | 0.59 | 0.65 | 0.49 | 0.66 | 0.53 | 0.74 | 0.71 | 0.79 | 30-Apr |
| TX | 0.59 | 0.66 | 0.71 | 0.69 | 0.65 | 0.66 | 0.67 | 0.61 | 0.58 | 30-Apr |
| UT | 0.64 | 0.76 | 0.64 | 0.66 | 0.64 | 0.69 | 0.60 | 0.59 | 0.63 | No Order |
| VA | 0.69 | 0.67 | 0.58 | 0.80 | 0.67 | 0.72 | 0.56 | 0.58 | 0.71 | 10-Jun |
| VT | 0.55 | 0.65 | 0.54 | 0.55 | 0.45 | 0.58 | 0.57 | 0.46 | 0.49 | 15-May |
| WA | 0.53 | 0.56 | 0.71 | 0.74 | 0.74 | 0.63 | 0.64 | 0.58 | 0.64 | No Expiration |
| WI | 0.62 | 0.69 | 0.64 | 0.69 | 0.64 | 0.66 | 0.60 | 0.63 | 0.60 | 13-May |
| WV | 0.56 | 0.57 | 0.57 | 0.62 | 0.63 | 0.72 | 0.55 | 0.59 | 0.61 | 3-May |
| WY | 0.49 | 0.54 | 0.41 | 0.56 | 0.43 | 0.61 | 0.55 | 0.61 | 0.43 | No Order |
| Total | 0.61 | 0.64 | 0.65 | 0.67 | 0.62 | 0.61 | 0.59 | 0.57 | 0.62 |  |

A review of the COVID-19 Reopening Readiness Index data reveals a steady decrease in New Jersey and Kansas, suggesting that those states are likely ready to ease some restrictions and move to the next phase of reopening (See Figure 1 for details). Vermont also displays signs that the spread of the COVID-19 has slowed. Hawaii shows signs of “flattening the curve” as the daily COVID-19 Reopening Readiness Index value has remained relatively stable for several consecutive days. Similarly, this behavior can also be seen for both Nebraska and Connecticut.

**Figure 1.**
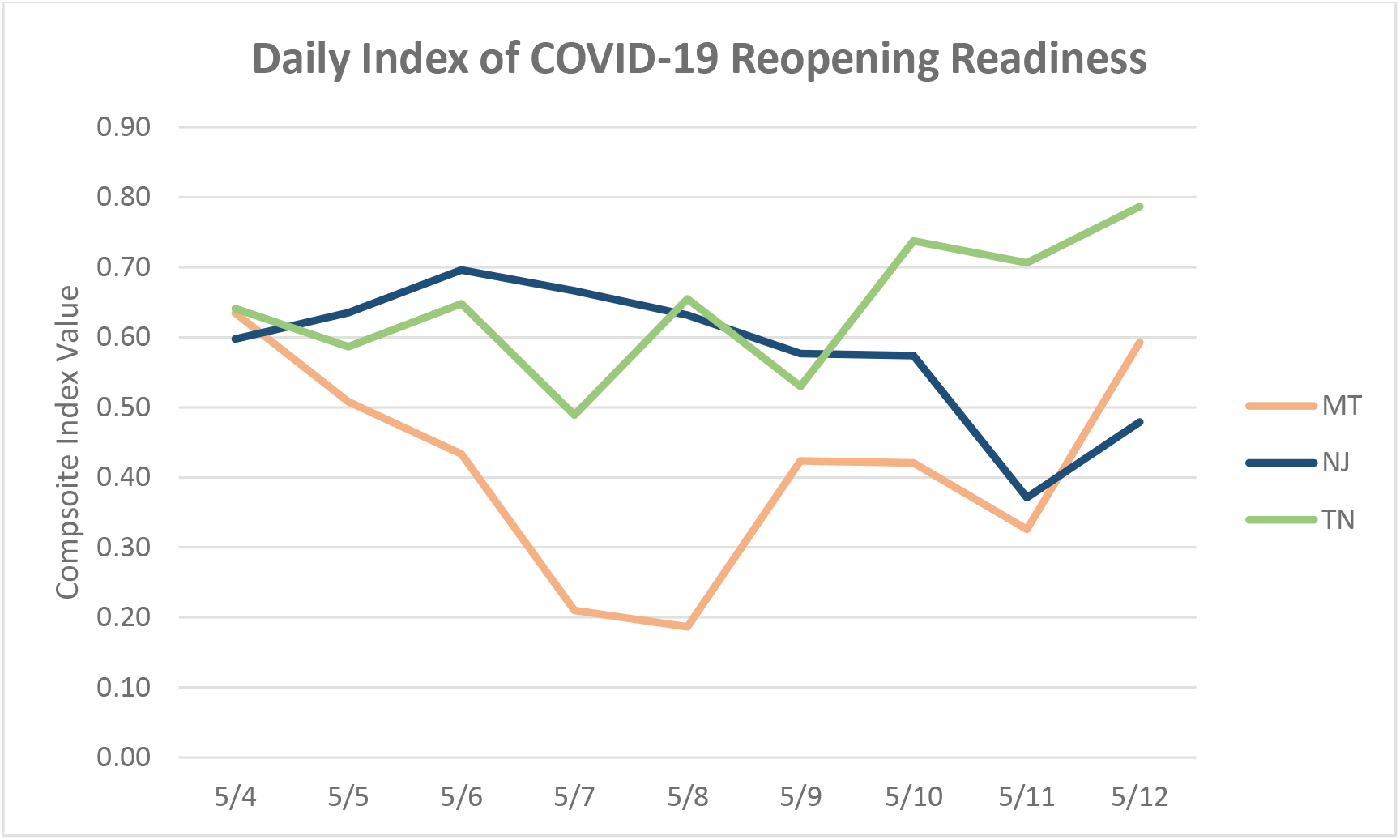
Exploration into Selected States

Conversely, a recent upward trend is observed in both Arkansas and Tennessee, requiring caution prior to moving to the next phase of reopening. A review of sub-indexes indicates a recent increase in the number of COVID-19 cases in both states. Georgia was one of first states that reopened its economy. While its COVID-19 Reopening Readiness Index value remained stable for the last three days, it shows little evidence of declining.

One thing to note is that the COVID-19 Reopening Readiness Index values for states such as Alaska, Montana and Kentucky exhibit greater volatility than states with a higher population mainly due to small or missing numbers as well as the delayed or irregular reporting of data observed in the data set. Caution is needed in interpreting the indexes of states with a relatively small population as index values can be fluctuating and misleading due to small daily counts.

## Index Deployment and Benefits

The COVID-19 Reopening Readiness Index provides a universal and unbiased perspective of viewing each state’s progress towards containing the spread of the virus or regressing over time. It can serve as an indicator of changes in the state’s readiness to reopen and move to the next phase. The following list illustrates some of the potential benefits of using the index:

> *Data driven, universal and unbiased:* The data-driven and standardized index enables a more consistent and transparent way of determining a state’s reopening readiness. This will ultimately contribute to reducing the overall rate of infection and facilitate the return to work and normalcy in general.
>
> *Key indicators for informed decision making:* States can determine if their restrictions are still needed and detect when there are adverse consequences of opening as well as if there is a need to revert to stricter rules.
>
> *Simple and timely:* The composite index minimizes the reliance on numerous measures provided by states, which can often vary and be misleading. It is one comprehensive measure that gives a summary of the status for each state. It is also easily interpretable which allows people to find patterns and identify trends in the data.
>
> *Comparison:* Because of its standardized view, states can be compared to each other. Every state can see the spread in other states and take counter measures if needed.
>
> *Business Application:* The index can also serve as a useful guide to any business dealing with the public and provide information regarding each state’s readiness to reopen and move to the next phase. Businesses can identify relative changes in the key underlying metrics over time using the reopening sub-indexes.
>
> *Healthcare needs:* The reopening index can act as an unbiased indicator to government and nongovernment healthcare agencies as to the anticipated demand for future healthcare services.

## Limitations, Future Study and Refinement

This analysis is merely the first step and will lay the foundation for follow-up analyses. This section discusses ideas to refine the current approach to calculating the COVID-19 Reopening Readiness Index and suggestions for future research.

> *Data Quality:* According to *The COVID Tracking Project*, the majority of data are directly from the state public health websites and some are derived from other sources. Additionally, definitions of some metrics might not be exactly the same, and the metrics are not always up to date. This can potentially undermine the accuracy and timeliness of the COVID-19 Reopening Readiness Index. Future research can address the delayed or irregular updates as well as possible data errors to improve the index accuracy as well as the data quality.
>
> *Other Metrics:* The COVID-19 Reopening Readiness Index is comprised of five sub-indexes, and the number of indicators used for the construction of sub-indexes is limited to five for simplicity, completeness and reliability of data. For example, not all states report the number of influenza-like illnesses or COVID-19-like syndromic cases. Therefore, other data sources and types of publicly available data relevant to each of the criterion relevant to COVID-19 (e.g. percent of ICU bed occupied, total tests per 100,000 people and the number of people recovered) can be considered to enhance the current index. It is also recommended that future research expand the index at the county and zip code level, break it down by demographic characteristics such as age group, and develop other customized indexes.
>
> *Index Calculation and Validation:* The three-day rolling average was used in this study to capture any recent trends that might have appeared in the data. Considering the irregular or delayed updates of data across data sources as well as missing or small daily counts often found in the data set, the five- or seven-day window can be explored. A longer rolling average will be more stable, but less responsive to the ever-changing situation. While standard deviations of the mean were used to substitute extreme values in favor of simplicity and for timely evaluation of the current pandemic trends, future research should consider more adequate treatment of outliers and missing data. Mean or median substitution can be also considered. Instead of equal weight given to each sub-index, different weights can be explored based on the importance of each index to generate a weighted average index number. Finally, the applicability of the index, whether it accurately reflects the current pandemic trends, needs to be validated based on the observed trends and/or using another data set.
>
> *Alternative to Index:* Our approach to the construction of the COVID-19 Reopening Readiness Index was easy-to-understand and simple-to-create to facilitate the understanding and interpretation of a state’s readiness status. A wide range of fluctuation in the daily COVID-19 counts however, would result in volatile index values, making trends difficult to evaluate. Therefore, alternative approaches such as the standard deviation showing the degree of variation from the mean can be considered. This would display whether the daily count falls within the expected range, making trend assessment easier.
>
> *Potential Uses for Business:* For businesses dealing with the public, the index can be used along with internal business metrics to generate much needed visibility into each state’s status. When it is visualized on a map, each state’s progress toward the next phase of reopening can be easily identified. External metrics such as the state’s unemployment rates, political party affiliation, and the current phase and details of reopening can be also developed. For example, external metrics to indicate the types of reopened business and the percent capacity can be used along with the COVID-19 Reopening Readiness Index and internal business measures to predict the demand for products or services.

## Data Availability

All of the data is available for public use.

## Appendix

**Table 3.** Sample Sub- and Composite Indexes for May 12.

| State | Mortality | Testing Capacity | Cases | Infection Rate | Hospitalization | Composite |
| --- | --- | --- | --- | --- | --- | --- |
| **AK** | 0.00 | 0.63 | 0.79 | 0.53 | 0.98 | **0.58** |
| **AL** | 0.71 | 0.76 | 0.59 | 0.84 | n/a | **0.73** |
| **AR** | 0.53 | 0.69 | 0.77 | 0.89 | 0.68 | **0.71** |
| **AZ** | 0.54 | 0.62 | 0.57 | 0.60 | 0.78 | **0.62** |
| **CA** | 0.41 | 0.66 | 0.56 | 0.67 | 0.75 | **0.61** |
| **CO** | 0.61 | 0.75 | 0.60 | 0.65 | 0.68 | **0.66** |
| **CT** | 0.37 | 0.56 | 0.63 | 0.62 | 0.67 | **0.57** |
| **DE** | 0.60 | 0.63 | 0.64 | 0.66 | 0.71 | **0.65** |
| **FL** | 0.43 | 0.63 | 0.68 | 0.68 | n/a | **0.61** |
| **GA** | 0.43 | 0.61 | 0.73 | 0.69 | n/a | **0.61** |
| **HI** | 0.00 | 0.40 | 1.00 | 0.71 | n/a | **0.53** |
| **IA** | 0.60 | 0.53 | 0.85 | 0.73 | 0.71 | **0.68** |
| **ID** | 0.00 | 0.66 | 0.67 | 0.79 | n/a | **0.53** |
| **IL** | 0.52 | 0.53 | 0.83 | 0.71 | 0.72 | **0.66** |
| **IN** | 0.43 | 0.73 | 0.59 | 0.68 | 0.69 | **0.62** |
| **KS** | 0.08 | 0.84 | 0.37 | 0.73 | n/a | **0.51** |
| **KY** | 0.59 | 0.16 | 0.79 | 0.51 | 0.78 | **0.56** |
| **LA** | 0.44 | 0.74 | 0.42 | 0.67 | 0.74 | **0.60** |
| **MA** | 0.33 | 0.74 | 0.52 | 0.72 | 0.72 | **0.61** |
| **MD** | 0.52 | 0.64 | 0.55 | 0.79 | 0.69 | **0.64** |
| **ME** | 0.23 | n/a | 0.49 | 0.91 | 0.54 | **0.54** |
| **MI** | 0.35 | 0.73 | 0.65 | 0.57 | 0.66 | **0.59** |
| **MN** | 0.44 | 0.67 | 0.63 | 0.85 | 0.78 | **0.67** |
| **MO** | 0.60 | 0.29 | 0.50 | 0.62 | 0.67 | **0.54** |
| **MS** | 0.62 | n/a | 0.57 | 0.49 | 0.74 | **0.61** |
| **MT** | 0.00 | n/a | 1.00 | 0.63 | 0.75 | **0.59** |
| **NC** | 0.65 | 0.71 | 0.53 | 0.76 | 0.67 | **0.66** |
| **ND** | 0.45 | 0.74 | 0.72 | 0.76 | 0.86 | **0.71** |
| **NE** | 0.45 | 0.75 | 0.41 | 0.75 | n/a | **0.59** |
| **NH** | 0.28 | n/a | 0.42 | 0.73 | 0.79 | **0.56** |
| **NJ** | 0.49 | 0.67 | 0.51 | 0.03 | 0.69 | **0.48** |
| **NM** | 0.43 | 0.67 | 0.67 | 0.67 | 0.77 | **0.64** |
| **NV** | 0.38 | 0.59 | 0.67 | 0.59 | 0.86 | **0.62** |
| **NY** | 0.43 | 0.74 | 0.51 | 0.69 | 0.66 | **0.61** |
| **OH** | 0.93 | 0.77 | 0.56 | 0.70 | 0.84 | **0.76** |
| **OK** | 0.45 | n/a | 0.81 | 0.93 | 0.81 | **0.75** |
| **OR** | 0.23 | 0.86 | 0.36 | 0.80 | 0.89 | **0.63** |
| **PA** | 0.46 | 0.66 | 0.58 | 0.75 | 0.71 | **0.63** |
| **RI** | 0.38 | 0.71 | 0.59 | 0.74 | 0.70 | **0.62** |
| **SC** | 0.75 | n/a | 0.49 | 0.58 | n/a | **0.61** |
| **SD** | 0.75 | 0.83 | 0.36 | 0.76 | 0.69 | **0.68** |
| **TN** | 1.00 | 0.65 | 0.77 | 0.73 | n/a | **0.79** |
| **TX** | 0.24 | 0.80 | 0.39 | 0.75 | 0.74 | **0.58** |
| **UT** | 0.45 | 0.74 | 0.47 | 0.76 | 0.71 | **0.63** |
| **VA** | 0.76 | n/a | 0.60 | 0.76 | 0.71 | **0.71** |
| **VT** | 0.00 | 0.71 | 0.54 | 0.61 | 0.60 | **0.49** |
| **WA** | 0.45 | 0.73 | 0.70 | 0.74 | 0.57 | **0.64** |
| **WI** | 0.36 | 0.68 | 0.51 | 0.72 | 0.71 | **0.60** |
| **WV** | 0.60 | 0.67 | 0.40 | 0.71 | 0.69 | **0.61** |
| **WY** | 0.00 | n/a | 0.72 | 0.40 | 0.60 | **0.43** |
| **Total** | 0.45 | 0.68 | 0.58 | 0.65 | 0.72 | **0.62** |

Note. n/a indicates no index available due to the absence of relevant data.

## References

Blackmon. D. (April 28, 2020). GOP States vs. Democrat States: The differences are very clear. Retrieved from https://dbdailyupdate.com/index.php/2020/04/28/gop-states-vs-democrat-states-the-differences-are-very-clear/

Brewster, J. (May 5, 2020). 87% of the states that have reopened voted for trump in 2016. Retrieved from Staffhttps://www.forbes.com/sites/jackbrewster/2020/05/05/87-of-the-states-that-have-reopened-voted-for-trump-in-2016/#4f9915ba242

Colorado Department of Public Health & Environment (CDPHE, 2020). Colorado COVID-19 updates data: Incidence & epidemic curve. Retrieved from https://covid19.colorado.gov/data/incidence-epidemic-curve

Fink, J. (April 14, 2020). What U.S. states plan to ease coronavirus lockdown restrictions? Retrieved from https://www.newsweek.com/what-us-states-plan-ease-coronavirus-lockdown-restrictions-1497802

NC Department of Health and Human Services (NCDHHS, 2020). COVID-19 North Carolina Dashboard. Retrieved from https://covid19.ncdhhs.gov/dashboard#key-metrics

The COVID Tracking Project (COVID Tracking, 2020). Retrieved from https://covidtracking.com/about-data/faq

The White House (2020). Guidelines opening up American again. Retrieved from https://www.whitehouse.gov/openingamerica/

